# Deep learning reveals diverging effects of altitude on aging

**DOI:** 10.1101/2024.09.25.24314218

**Authors:** Amanuel Abraha Teklu, Indra Heckenbach, Michael Angelo Petr, Daniela Bakula, Guido Keijzers, Morten Scheibye-Knudsen

## Abstract

Aging is influenced by a complex interplay of multifarious factors, including an individual’s genetics, environment, and lifestyle. Notably, high altitude may impact aging and age-related diseases through exposures such as hypoxia and ultraviolet radiation. To investigate this, we mined summary exposure value as a measure of risk exposure levels, and disability-adjusted life years (DALYs) as a measure of disease burden from the Global Health Data Exchange (GHDx) for each subnational region of Ethiopia, a country with considerable differences in the living altitude. We conducted a cross-sectional clinical trial involving 227 highland and 202 lowland dwellers from the Tigray region in Northern Ethiopia to gain a general insight into the biological aging at high altitudes. Notably, we observed significantly lower risk exposure rates and a reduced disease burden in higher-altitude regions of Ethiopia. When assessing biological aging using facial photographs, we found a faster rate of aging with increasing elevation, likely due to greater UV exposure. Conversely, analysis of nuclear morphologies of peripheral blood mononuclear cells in blood smears (PBMCs) with five different senescence predictors revealed a significant decrease in DNA damage-induced senescence in both monocytes and lymphocytes with increasing elevation. Overall, our findings suggest that disease and DNA damage-induced senescence decreases with altitude in agreement with the idea that oxidative stress may drive aging.

## Introduction

Aging and age-related diseases are critically regulated by multifarious factors pertained to the genetics, metabolism, behavior, lifestyle, and environment of the individual or population ^1^. The role of oxidative stress in aging, particularly how reduced or increased oxygen exposure affects aging remains a debated topic ^2^. At high altitudes, hypoxia occurs due to a decrease in the partial pressure of oxygen in the air, accompanied by increased exposure to ultraviolet (UV) radiation ^3^. While oxygen is essential for cellular energy production in the form of ATP through oxidative phosphorylation in the mitochondria, it is also highly reactive and can oxidize proteins, lipids, DNA, and other macromolecules in cells. Data suggests that cells incubated under hypoxic condition may exhibit increased genetic instability ^4^, altered metabolism ^5^ and cell death activities ^6^ as well as therapy resistant behaviors ^7^. Nevertheless, there are emerging studies demonstrating the beneficial role of hypoxia in health and aging. For instance, culturing primary cells in low oxygen can improve cell survival ^8^ and hypoxia is required for maintaining the stemness of stem cells ^9^. Further, reduced oxygen levels have been implicated in the considerable longevity of naked mole rats which reside in hypoxic underground tunnels and live longer, healthier lives compared to similarly sized rodents in normoxic environments ^10^. In addition, nematodes exposed to moderate hypoxia have shown extended lifespan by up to 40 percent ^11^.

However, there are currently no aging studies on native human population living at high-altitude hypoxia to validate these findings. Investigating the impact of hypoxia on human aging is challenging due to the limited number of populations living at both high and low altitudes. Therefore, Ethiopia is of particular interest because it contains the highest proportion of population in the world permanently living at high altitude (> 1500 masl) ^12^. At the organismal level, the body undergoes adaptation at high altitude, where tissues consume reduced oxygen by entering into a hypometabolic state to adjust to the low level of oxygen at high altitudes ^13^. In addition, recent studies have also identified nitric oxide (NO) and cyclic guanosine monophosphate (cGMP) mediated vasodilation as a mechanism of adaptation to high altitude chronic hypoxia ^14^. Although organisms employ various adaptative mechanisms to hypoxia and other stimuli to maintain homeostasis, emerging evidence suggests these mechanisms decline with age ^15^. For example, aged mice show a reduced HIF1 function compared to younger mice ^16^. In addition, in a study conducted on aged human diploid fibroblasts, a significant reduction in HIF1 binding efficiency to the sequences of hypoxia response elements (HREs) was observed ^17^. Furthermore, as age advances, oxygen sensing ^18^ and the ventilatory system of the body ^19^ also changes, potentially leading to a relative hypoxia. Weakening of the respiratory system combined with the failure of the adaptation machinery to hypoxia with age may increase the vulnerability of older individuals, especially those dwelling at higher altitudes.

Taken together, existing *in vitro*, data, animal models, and human studies suggests that high altitude may influence human aging ^10,17,20,21^. To test this hypothesis, we initially assessed the risk factors and disease burden in lowland and highland regions of Ethiopia using summary exposure value (SEV) and disability-adjusted life years (DALYs) data from the Global Health Data Exchange (GHDx), respectively. We then cross-sectionally collected biomarkers (facial image and nuclear images of immune cell from peripheral blood) from a cohort of 429 human volunteers dwelling at varying altitudes (highlands and lowlands) in the Tigray region of Norther Ethiopia. Using computational approaches, we examined whether these biomarkers of biological aging were altered in response to altitude. Overall, this study provides new insights into the rate of biological aging and the levels of risk factors and diseases burden at high altitudes in a low-income country, potentially guiding future mechanistic and intervention studies on aging in high-altitude environments.

## Result

### Lower risk exposure levels at higher altitude regions of Ethiopia

To investigate how regional elevation (Fig S1) might impact the exposome, we mined estimated risk exposure values on all risk factors and on 84 individual risk factors in the form of age-standardized summary exposure value (SEV) rates for the nine regions and two chartered cities of Ethiopia from the GHDx of the Institute for Health Metrics and Evaluation (IHME). Findings from the GBD 2019 Ethiopia subnational analysis showed the presence of a significant difference in socio-demographic index (SDI) among the Ethiopian regions, and this might impact their risk exposure levels as well. We therefore normalized the SEV values of each region by their corresponding SDI. The normalized SEV rates of each region was then correlated with their corresponding average elevation to assess the impact. As a result, we found a significantly lower rate of age-adjusted SDI normalized summary exposure values for all risk factor (r = - 0.74, p = 0.01) as the elevation of the regions increases, indicating overall risk factor level and exposure are lower at higher altitude regions (Fig 1a). Similar trends were also observed for level 1 risk factors including behavioral (r = - 0.80, p = 0.003), environmental (r = - 0.70, p = 0.02), and metabolic (r = - 0.34, p = 0.31) risk factors (Fig 1b-d). In contrast, nitrogen dioxide pollution (r = 0.62, p = 0.04), alcohol use (r = 0.12, p = 0.73) and ambient particulate matter pollution (r = 0.39, p = 0.24) has shown increasement at higher altitude regions. Results from correlation of all types of risk factors is found in supplemental table 1.

**Figure 1:**
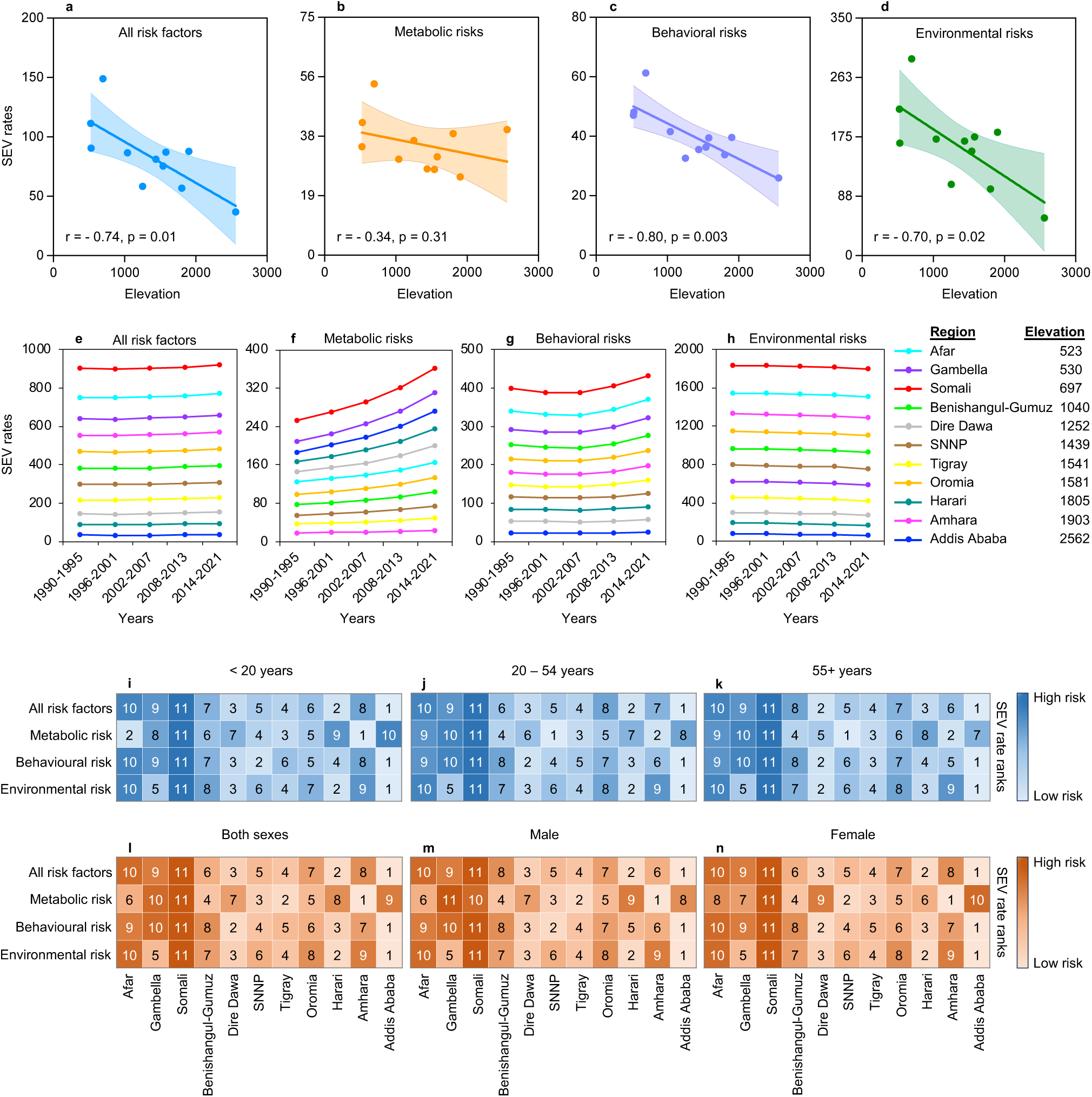
Elevation based regional variation in risk exposure levels in Ethiopia. a. Scatter plot of SEV rates of all risk factors with mean elevation of the subnational regions of Ethiopia. b. Scatter plot of SEV rates of metabolic risks with mean elevation of the subnational regions of Ethiopia. c. Scatter plot of SEV rates of behavioral risks with mean elevation of the subnational regions of Ethiopia. d. Scatter plot of SEV rates of environmental risks with mean elevation of the subnational regions of Ethiopia. e. Trends of SEV rates of all risk factors in each subnational region of Ethiopia from 1990 to 2021. f. Trends of SEV rates of metabolic risks in each subnational region of Ethiopia from 1990 to 2021. g. Trends of SEV rates of behavioral risks in each subnational region of Ethiopia from 1990 to 2021. h. Trends of SEV rates of environmental risks in each subnational region of Ethiopia from 1990 to 2021. i. Heatmap of ranks of SEV rates of all risk factors, metabolic, behavioral and environmental risks in under 20 years old population in each subnational region of Ethiopia in 2021. j. Heatmap of ranks of SEV rates of all risk factors, metabolic, behavioral and environmental risks in 20 to 54 years old population in each subnational region of Ethiopia in 2021. k. Heatmap of ranks of SEV rates of all risk factors, metabolic, behavioral and environmental risks in above 54 year old population in each subnational region of Ethiopia in 2021. l. Heatmap of ranks of SEV rates of all risk factors, metabolic, behavioral and environmental risks in both sexes in each subnational region of Ethiopia in 2021. m. Heatmap of ranks of SEV rates of all risk factors, metabolic, behavioral and environmental risks in males in each subnational region of Ethiopia in 2021. n. Heatmap of ranks of SEV rates of all risk factors, metabolic, behavioral and environmental risks in females in each subnational region of Ethiopia in 2021.

### Lower rates of disease burden are observed at higher altitude regions of Ethiopia

Regional variation in disease burden and injuries attributed to different altitude levels remained poorly studied. Here we collected rates of age-adjusted disability-adjusted life years (DALYs) as a measure of disease burden from GHDx of the IHME for the 11 subnational regions of Ethiopia. Estimated DALYs values were obtained for all cause as well as for 370 distinct causes of disease burden (disease types). We also obtained age-adjusted DALYs rates for each region for the 92 different diseases which the GBD 2017 study on population aging identified as age-related ^22^. The effect of regional elevation on disease burden were assessed by correlating the age-adjusted SDI normalized DALYs rates of each region with their average elevation. Our analysis showed a significant decrease in the age-adjusted SDI normalized DALY rates with elevation for all cause combined (r = - 0.76, p = 0.007) (Fig 2a) as well as for all of the level 1 causes including non-communicable diseases (r = - 0.73, p = 0.01), communicable disease (r = - 0.75, p = 0.008) and injuries (r = - 0.71, p = 0.01) (Fig 2b-d). In contrary, Schistosomiasis (r = 0.03, p = 0.92) and leishmaniasis (r = 0.21, p = 0.54) has shown an increased trend with elevation although the relationship was not significant (Supplemental Table 2). Similarly, significantly decreased DALY rates were found for most of the age-related diseases at higher altitude regions of Ethiopia (Supplemental Table 3). In sum, these findings could suggest that high altitude may reduce the pace of aging.

**Figure 2:**
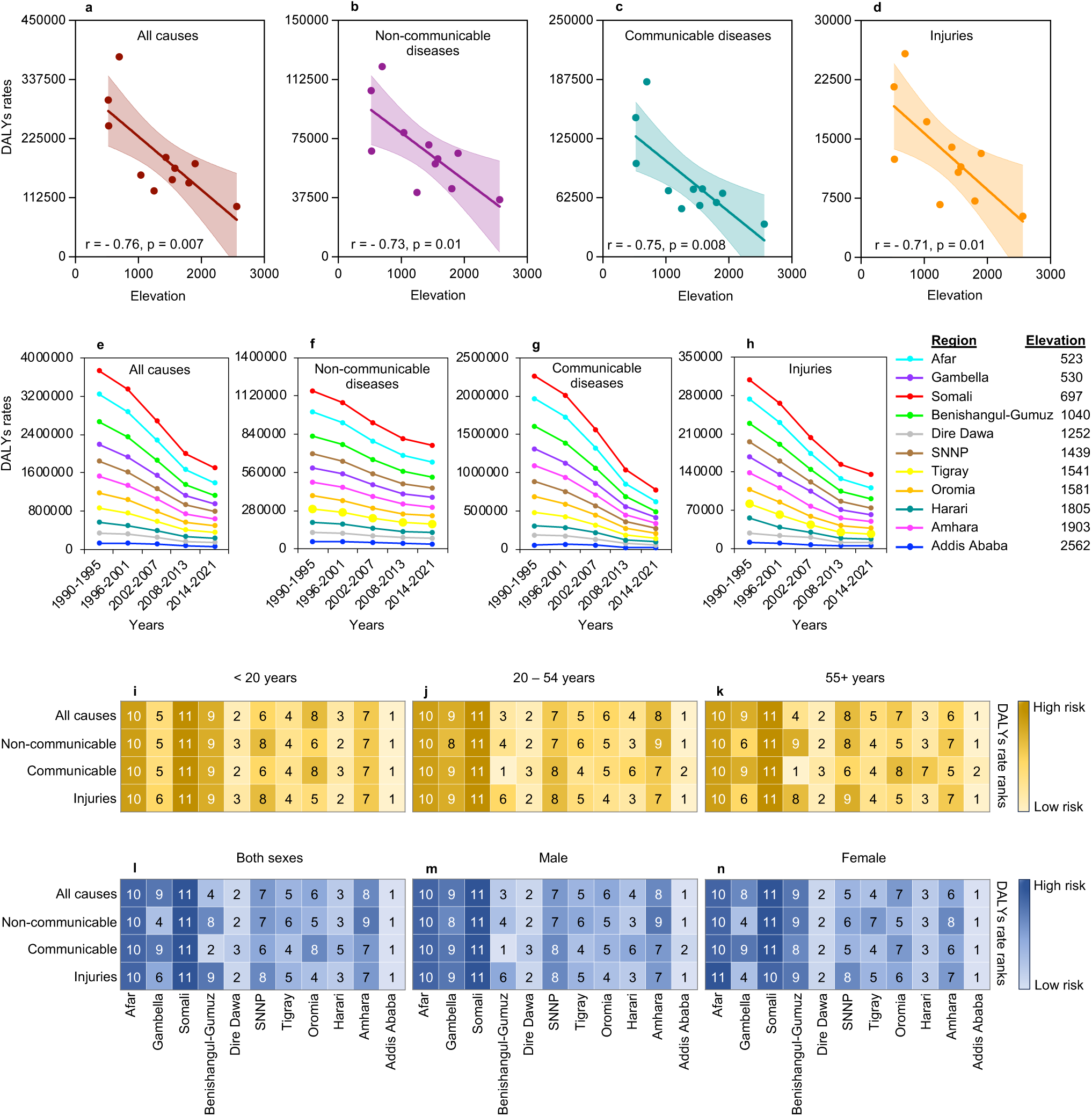
Regional altitude-induced variation in disease burden in Ethiopia. a. Scatter plot of DALYs rates of all causes with mean elevation of the subnational regions of Ethiopia. b. Scatter plot of DALYs rates of non-communicable diseases with mean elevation of the subnational regions of Ethiopia. c. Scatter plot of DALYs rates of communicable diseases with mean elevation of the subnational regions of Ethiopia. d. Scatter plot of DALYs rates of injuries with mean elevation of the subnational regions of Ethiopia. e. Trends of DALYs rates of all causes in each subnational region of Ethiopia from 1990 to 2021. f. Trends of DALYs rates of non-communicable diseases in each subnational region of Ethiopia from 1990 to 2021. g. Trends of DALYs rates of communicable diseases in each subnational region of Ethiopia from 1990 to 2021. h. Trends of DALYs rates of injuries in each subnational region of Ethiopia from 1990 to 2021. i. Heatmap of ranks of DALYs rates of all causes, non-communicable, communicable and injuries in under 20 years old population in each subnational region of Ethiopia in 2021. j. Heatmap of ranks of DALYs rates of all causes, non-communicable, communicable and injuries in 20 to 54 years old population in each subnational region of Ethiopia in 2021. k. Heatmap of ranks of DALYs rates of all causes, non-communicable, communicable and injuries in above 54 years old population in each subnational region of Ethiopia in 2021. l. Heatmap of ranks of DALYs rates of all cause, non-communicable, communicable and injuries in both sexes in each subnational region of Ethiopia in 2021. m. Heatmap of ranks of DALYs rates of all causes, non-communicable, communicable and injuries in males in each subnational region of Ethiopia in 2021. n. Heatmap of ranks of DALYs rates of all causes, non-communicable, communicable and injuries in females in each subnational region of Ethiopia in 2021.

### Increased photoaging is observed at higher living altitudes

To gain insight into whether a varied biological aging is observed in response to exposure to high altitude chronic hypoxia and other related factors, we investigated aging biomarkers of the highland and lowland dwellers in the Tigray region in Northern Ethiopia (Fig S3). Given the challenging conditions of working in the less developed areas, biomarkers were chosen that could be collected with relative ease. Previous studies have suggested that facial images can be used as a biomarker to estimate biological aging ^31^. Based on this, we cross sectionally collected facial images from a cohort of 429 participants dwelling at varying altitudes (600 to 3200 masl) in the Tigray region of Northern Ethiopia (Table 1). We applied four different deep learning algorithms (Fig S3) to these images and generated predicted age for each participant using the three algorithms that had the most accurate predictive age-association (Fig S4). The age difference calculated by subtracting the chronological age from the predicted age were correlated with the average residential elevation of each participant, and we found that the rate of facial aging generally increases as the living altitude of the participants increases (r = 0.14, p = 0.0027) (Fig 3b). However, a decreased aging rate was shown as the chronological age of the participants increases (r = - 0.20, p < 0.0001) (Fig 3c). We also investigated the impact of body mass index, sex, and residence on the rate of aging of the participants, and no significant effect of these factors was found on the rate of aging of the participants (Fig 3d-f).

**Table 1.** Covariate information of the study participants

| Characteristics | Lowland |  |  |  | Highland |  |  |  |  | P value |
| --- | --- | --- | --- | --- | --- | --- | --- | --- | --- | --- |
| Study sites | Sheraro<br>(n=105) | Dedebit<br>(n=50) | Alamata<br>(n=47) | Combined<br>(n=202) | Hagereselam<br>(n=51) | Edagahamus<br>(n=50) | Korem<br>(n=54) | Maychew<br>(n=72) | Combined<br>(n=227) |  |
| Average elevation(masl) | 988 | 912 | 1544 | 1093 | 2534 | 2689 | 2446 | 2481 | 2537 | < 0.001 |
| Chronological age | 31 ± 18 | 37 ± 24 | 37 ± 17 | 34 ± 20 | 31 ± 14 | 36 ± 18 | 27 ± 16 | 40 ± 24 | 33 ± 19 | 0.60 |
| Sex |  |  |  |  |  |  |  |  |  |  |
| Female | 50 | 21 | 25 | 96 | 24 | 23 | 28 | 31 | 106 |  |
| Male | 55 | 29 | 22 | 106 | 27 | 27 | 26 | 41 | 121 |  |
| Residence |  |  |  |  |  |  |  |  |  |  |
| Urban | 75 | 0 | 37 | 112 | 31 | 33 | 33 | 50 | 147 |  |
| Rural | 30 | 50 | 10 | 90 | 20 | 17 | 21 | 22 | 80 |  |
| Height (m) | 155 ± 23 | 158 ± 15 | 165 ± 8 | 158 ± 19 | 161 ± 17 | 158 ± 16 | 152 ± 16 | 159 ± 14 | 157 ± 16 | 0.49 |
| Weight (kg) | 49 ± 15 | 47 ± 14 | 54 ± 10 | 50 ± 14 | 48 ± 11 | 49 ± 13 | 45 ± 13 | 50 ± 14 | 48 ± 13 | 0.23 |
| Body mass index (kg/m<sup>2</sup>) | 19 ± 3 | 19 ± 3 | 20 ± 3 | 19 ± 3 | 19 ± 3 | 19 ± 3 | 19 ± 3 | 20 ± 4 | 19 ± 3 | 0.57 |

**Figure 3:**
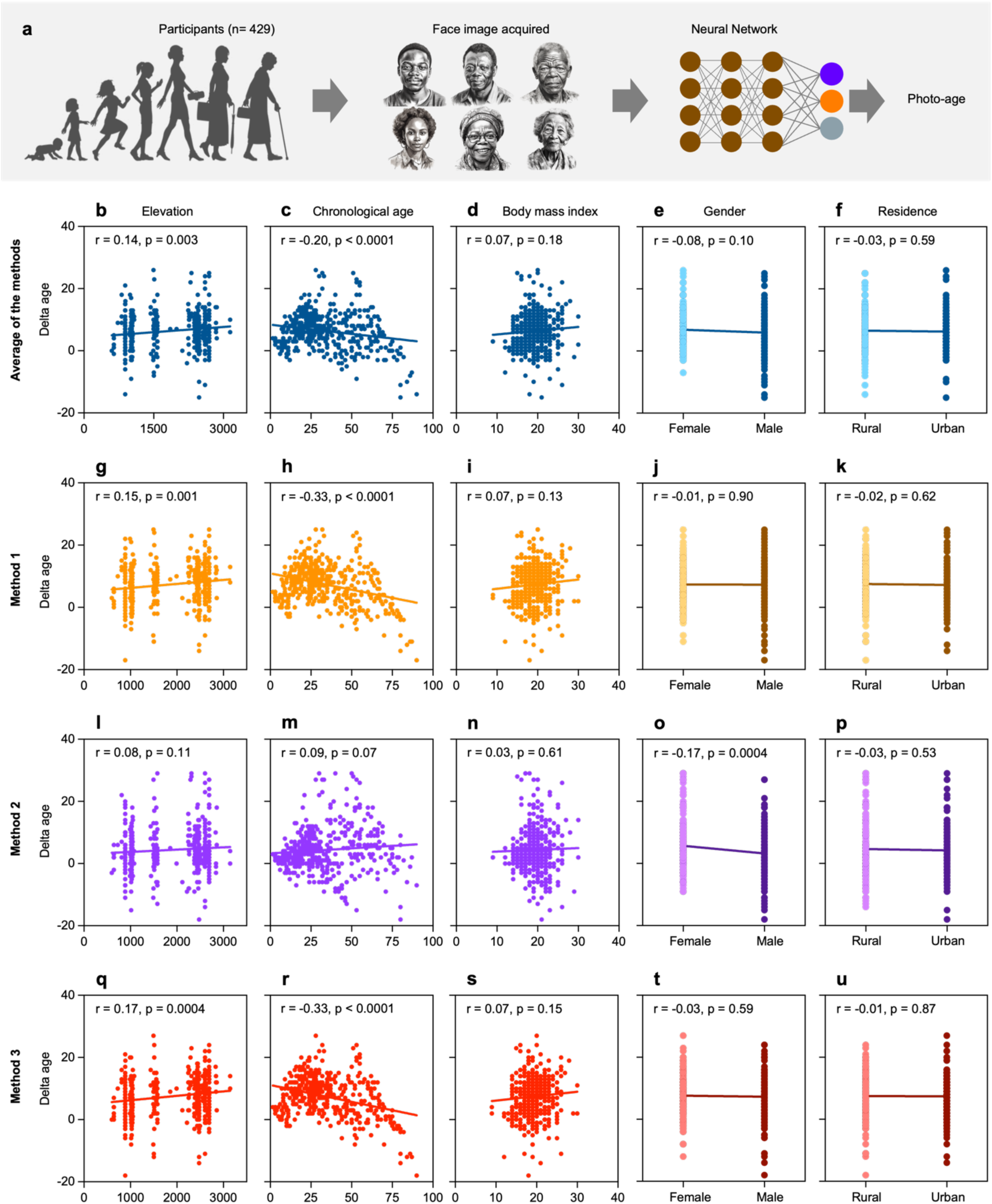
Rate of biological aging at different elevation levels in Tigray. a. Workflow showing age prediction from facial images of the lowland and highland participants. b. Scatter plot of average delta age with residential elevation of the study participants. c. Scatter plot of average delta age with chronological age of the participants. d. Scatter plot of average delta age with the body mass index of the study participants. e. Scatter plot of average delta age with the gender of the study participants. f. Scatter plot of average delta age with the residence of the study participants. g. Scatter plot of delta age of method 1 with the residential elevation of the participants. h. Scatter plot of delta age of method 1 with the chronological age of the participants. i. Scatter plot of delta age of method 1 with the body mass index of the participants. j. Scatter plot of delta age of method 1 with the gender of the participants. k. Scatter plot of delta age of method 1 with the residence of the participants. l. Scatter plot of delta age of method 2 with the residential elevation of the participants. m. Scatter plot of delta age of method 2 with the chronological age of the participants. n. Scatter plot of delta age of method 2 with the body mass index of the participants. o. Scatter plot of delta age of method 2 with the gender of the participants. p. Scatter plot of delta age of method 2 with the residence of the participants. q. Scatter plot of delta age of method 3 with the residential elevation of the participants. r. Scatter plot of delta age of method 3 with the chronological age of the participants. s. Scatter plot of delta age of method 3 with the body mass index of the participants. t. Scatter plot of delta age of method 3 with the gender of the participants. u. Scatter plot of delta age of method 3 with the residence of the participants.

### Reduced predicted DNA damage induced senescence was observed at higher altitudes

Nuclear morphology has been shown to be a biomarker of senescence ^23^. Thus, along with the facial images, we also collected images of the nucleus of immune cells using thin-layer blood smears from the cohort of 429 participants. To investigate senescence, we identified nucleated cells in the blood smears of the highland and lowland dwellers and then applied the nuclear senescence predictor (NUSP) of the highland and lowland dwellers (Fig 4a). We used five different NUSP models and generated senescence scores to assess the effect of high altitude on organismal aging at the cellular level (Fig 4b-u). Notably, our analysis showed significantly higher senescence scores with elevation using the replicative exhaustion (RS) for both monocytes (r = 0.21, p < 0.0001) and lymphocytes (r = 0.31, p < 0.0001) (Fig 4b), but significantly lower senescence scores for monocyte (r = - 0.25, p < 0.0001) using the ionizing radiation (IR) model (Fig 4c) as well as monocytes (r = - 0.15, p = 0.002) and lymphocytes (r = - 0.41, p < 0.0001) using the doxorubicin (Doxo) model (Fig 4d). In addition, increasing chronological age was significantly negatively correlated with the senescence scores detected by the IR model (monocytes: r = - 0.12, p = 0.01; lymphocytes: r = - 0.09, p = 0.049) (Fig 4h), but significantly positively correlated with the senescence scores from the Anti model (monocytes: r = 0.10, p = 0.04; lymphocytes: r = 0.11, p = 0.02) (Fig 4k). Examining the effect of increasing predicted age, we found lower scores for the senescence of monocytes detected by the IR model (r = - 0.14, p = 0.004) (Fig 4m), whereas higher scores for the senescence of lymphocytes detected by the Anti model (r = 0.11, p = 0.03) (Fig 4p). Rate of aging of the participants seems less important to affect monocyte senescence, but for lymphocytes, the Doxo model showed lower scores as the rate of aging of the participants increases (r = - 0.12, p = 0.02) (Fig 4s). Further more, we have investigated how the senescence status of the PBMCs is affected by the body mass index, sex, and rural or urban residence of the participants (Fig S5). As a result, detection by the Atvr (mitochondrial dysfunction) model showed lower senescence scores of monocytes in the participants with higher body mass index (r = - 0.12, p = 0.03) (Fig S5j).

**Figure 4:**
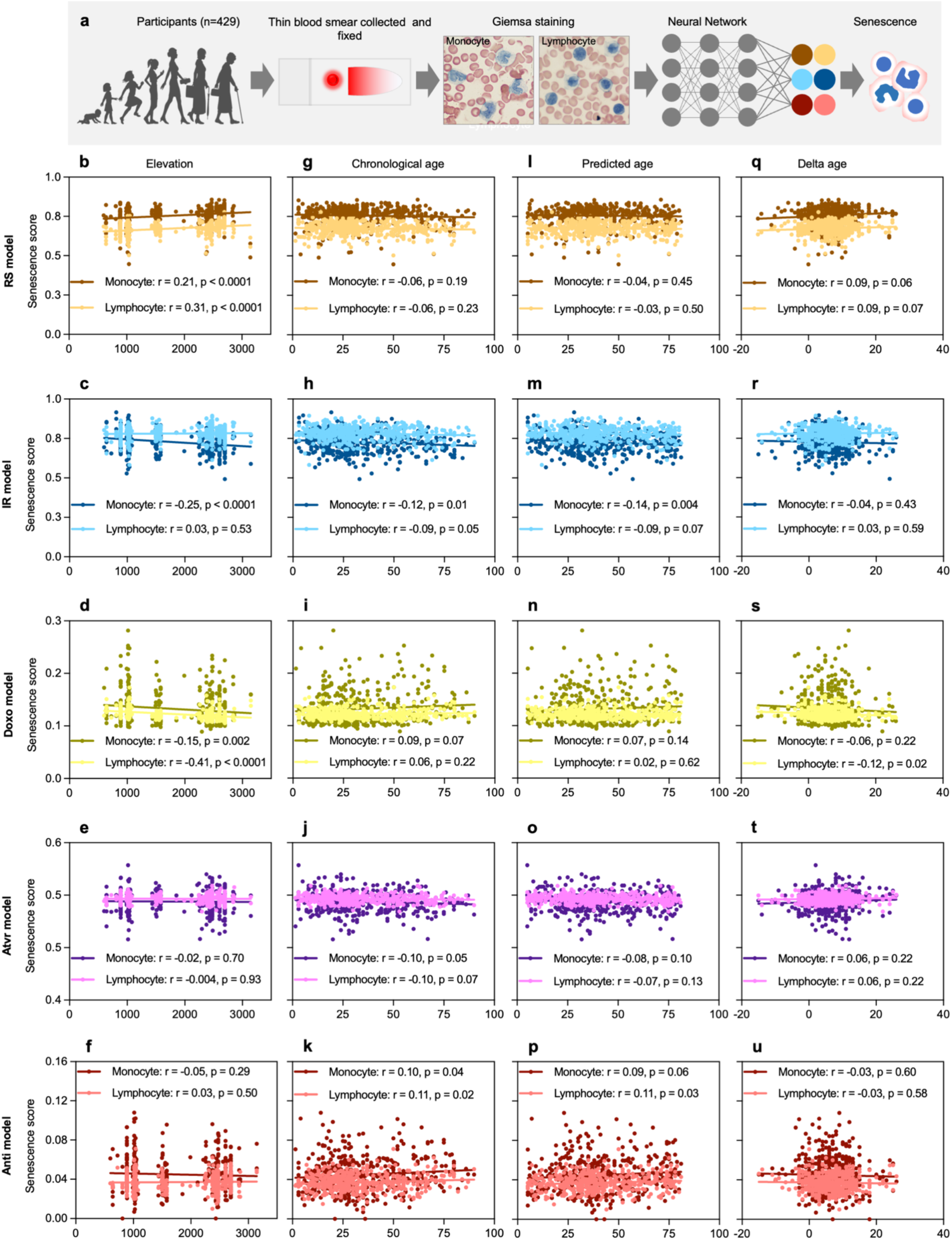
Senescence status of highland and lowland dwellers. a. Workflow of senescence prediction from nuclear morphology of PBMCs. b. Scatter plot of predicted senescence (RS model) of monocytes and lymphocytes with the residential elevation of the study participants. c. Scatter plot of predicted senescence (RS model) of monocytes and lymphocytes with the chronological age of the study participants. d. Scatter plot of predicted senescence (RS model) of monocytes and lymphocytes with the predicted age of the study participants. e. Scatter plot of predicted senescence (RS model) of monocytes and lymphocytes with the delta age of the study participants. f. Scatter plot of predicted senescence (IR model) of monocytes and lymphocytes with the residential elevation of the study participants. g. Scatter plot of predicted senescence (IR model) of monocytes and lymphocytes with the chronological age of the study participants. h. Scatter plot of predicted senescence (IR model) of monocytes and lymphocytes with the predicted age of the study participants. i. Scatter plot of predicted senescence (IR model) of monocytes and lymphocytes with the delta age of the study participants. j. Scatter plot of predicted senescence (Doxo model) of monocytes and lymphocytes with the residential elevation of the study participants. k. Scatter plot of predicted senescence (Doxo model) of monocytes and lymphocytes with the chronological age of the study participants. l. Scatter plot of predicted senescence (Doxo model) of monocytes and lymphocytes with the predicted age of the study participants. m. Scatter plot of predicted senescence (Doxo model) of monocytes and lymphocytes with the delta age of the study participants. n. Scatter plot of predicted senescence (Atvr model) of monocytes and lymphocytes with the residential elevation of the study participants. o. Scatter plot of predicted senescence (Atvr model) of monocytes and lymphocytes with the chronological age of the study participants. p. Scatter plot of predicted senescence (Atvr model) of monocytes and lymphocytes with the predicted age of the study participants. q. Scatter plot of predicted senescence (Atvr model) of monocytes and lymphocytes with the delta age of the study participants. r. Scatter plot of predicted senescence (Anti model) of monocytes and lymphocytes with the residential elevation of the study participants. s. Scatter plot of predicted senescence (Anti model) of monocytes and lymphocytes with the chronological age of the study participants. t. Scatter plot of predicted senescence (Anti model) of monocytes and lymphocytes with the predicted age of the study participants. u. Scatter plot of predicted senescence (Anti model) of monocytes and lymphocytes with the delta age of the study participants.

## Conclusion

Alterations in oxygen levels have been associated with health and disease ^25^. Our analysis revealed that increased altitude affects a wide range of risk factors and diseases as well as rate of biological aging. Notably, we saw a strong decrease in disability adjusted life-years suggesting that increased altitude is associated with better health. This supports previous findings that people at highlands live longer and often have better health than those living at lowlands ^26^. Other studies have, however, found an association of high altitude with higher prevalence of cancer and cancer attributable death rates ^27^ ^28^, in particular skin cancer ^29,30^. This is in agreement with our deep learning investigations on facial images that showed a faster rate of aging of highland dwellers than the lowland dwellers, and this is likely attributed to the high level of ultraviolet radiation at high altitude ^31,32^. Given the role of oxygen on driving oxidative stress we speculated that increased altitude might affect senescence. Accordingly, we found a reduction in DNA damage- related senescence (IR and Doxo), suggesting a protective role of high altitude in some aspects of biological aging. Surprisingly, we saw increased replicative associated senescence with increasing altitude suggesting that there may be phenotypical overlap between previously described hypoxia-induced senescence and replicative senescence. Accordingly, chronic hypoxia in sleep apnea patients leads to shortened telomeres ^33^. Taken together, our analyses showed that high altitude may be a key factor in modulating not only aging-associated risk factors and diseases but also biological aging. Overall, our study represents a novel effort in understanding the biological aging of the unique population of high-altitude residents of a low income country. Nurturing the efforts to understand the mechanism of survival and aging process at high altitude would generate knowledge that may benefit the global population in the prevention and treatment of disabilities and illnesses.

Our study has several limitations. For instance, to study the levels of risk factors and disease burden at different altitudes in Ethiopia, we used data from the Institute for Health Metrics and Evaluation. In Ethiopia, although currently improving, the data reporting system in some areas is still not technologically supported, and in such circumstances, the reporting is done manually, making it prone to mis- or over reporting or data loss leading to data bias. Further, in our study we only involved volunteers who are healthy and permanent or long-term residents in their territory. The healthy status of each participant was based on their declaration and based on the physical examination performed by the licensed health professional, and this may not reflect the biological situation of the participants. The residence history of the participants was also based on what they reported, and to minimize effects of changing residence from low to highland or vice versa. Our biomarker investigations have been done using largely noninvasive methodologies. Consequently, the limitations of these systems may have affected our results as well. We tried to minimize such effects by combining different methods to produce averaged data for analysis and presentation. In fact, a strong association is observed among predicted age by the different age predictor tools we used, suggesting low variation by methods. Further, senescence scores of each highland and lowland participant were deduced from stained nuclear morphology of the immune cells.

Despite color normalization, differences in staining intensities could subtly affect the generated senescence scores. To avoid such variation, we performed the staining of all smears of all participants simultaneously. In addition, during analysis, intensity normalization was performed at the individual level and between the lowland and highlanders. It is also important to underline that the microscopic identification of lymphocytes and monocytes from other blood cells in the smears was performed manually and based on cytoplasmic and nuclear characteristics, indicating the possibility of involving other cell that morphologically resemble monocyte or lymphocyte resembling cells in the analysis. To minimize this, the detection and capturing of monocytes and lymphocytes from the thin blood smear was performed by a licensed Medical Laboratory Technologist. It is also possible that the deep learning algorithms may have reduced accuracy in assessing senescence in blood smears. Nevertheless, it is relatively striking that two different predictors describe decreased DNA damage induced senescence with elevation.

Overall, our study has provided a comprehensive information on risk factors, diseases, and biological aging at high altitude suggesting that high altitude is associated with lower disease incidence and DNA damage induced senescence in blood. Our study also demonstrated the value of deep learning tools in research in areas where research facilities are limited such as in Ethiopia. And our works has provided pioneering insight about the population aging of a low-income country in Africa.

## Methods

### Study area and setting

Ethiopia is an East African country with an economic and social development classified among the least developed countries (LDCs) ^34^. Being the second most populous nation in Africa (estimated 109.5 million in 2024) ^35,36^, Ethiopia represents the highest proportion of native highland dwellers in the world ^12^. In chronological perspective, the Ethiopian population is young (median age 18.9) and the elderly population (65 years and above) accounted for 5.1% in 2010 and estimated to reach 10.3% by 2050 ^37^. The Federal Democratic Republic of Ethiopia has been administratively divided into 11 subnational regions, namely, Tigray, Amhara, Afar, Somali, Harar, Dire Dawa, Oromia, Addis Ababa, Gambella, Benshangul Gumuz, and South Nations Nationalities and People (SNNP). Interestingly, these subnational regions have highly varied mean elevation (Fig S1a-b), making it very suitable for investigating the impact of regional variation in elevation on risk exposure levels and disease burden. Further, within specific region such as Tigray, there exists a large difference in living altitude (600 to 3200 masl) (Fig S1c-d). Consequently, we chose the Tigray region to examine the impact of high altitude on human aging. Besides, the Tigray region has millions of people permanently dwelling at both higher and lower altitudes (Fig S2a-g), and these lowland and highland populations have similar sociodemographic index ^38^. To gain the most possible impact of high altitude on human aging, we selected participants from residential areas with the highest mean elevation in Tigray including Hagereselam, Edagahamus, Korem and Maychew. Sheraro, Dedebit and Alamata are among the lowland residential areas in Tigray.

### Data source for risk exposure and disease burden assessments

To investigate elevation based regional variation in risk exposure levels and disease burden in Ethiopia, estimated rate of age-adjusted summary exposure value (SEV) as a measure of risk exposure, and estimated rate of age-adjusted disability-adjusted life years (DALYs) as a measure of disease burden were minded from the Global Health Data Exchange (GHDx) of the Institute for Health Metrics and Evaluation^39^ for each of the 11 subnational regions (nine regions and two chartered cities) of Ethiopia for the year 2021. SEV data was collected for all risk factors as well as for level 1, level 2, level 3 and level 4 risk factors for each region.

Similarly, DALYs data was obtained for all cause, level 1, level 2, level 3 and level 4 causes for each region. The mean elevation of each region was determined and provided by the Tigray Statistical Agency (Tigray, Ethiopia). Data about the socio-demographic index (SDI) of each region was gathered from the GBD 2019 Ethiopia subnational analysis ^40^.

### Human participants for biological aging studies

We conducted a cross sectional clinical trial to examine the biological aging of highland and lowland populations in the Tigray region of Northern Ethiopia (ethical approval number: MU- IRB 2020/2022, see below). 429 volunteer participants were recruited, of which 53% (229) were native highland dwellers and 202 were lowland dwellers. All participation was voluntary and after informed consent provided. From all participants, we collected covariate data, facial images, and thin blood smears from April to May 2022.

### Sociodemographic data collection

We collected demographic data regarding the chronological age, sex, residence, ethnicity, movement history and health status of the study participants using a self-developed short questionnaire. The height and weight of each participant were recorded from stadiometer and digital scale, respectively. Body mass index (BMI) was calculated using the measured height and weight of each participant as previously described ^41^. Elevation data of the residential area (locally known as ‘‘Tabiya’’) of each participant was obtained from the Tigray Statistical Agency.

### Inclusion criteria

We involved residents who voluntarily provided their informed consent to participate in this study. In addition, it was a requirement that the volunteer was a permanent resident in a given place or moved to another place within Tigray, but with the same elevation. This was to ensure that the highland participants were not exposed to normoxia, or the lowland participants were not exposed to high-altitude hypoxia. Further, we considered only those volunteers who have self-declared that they have been healthy for the previous six months and had no finding on physical examination by the healthcare professional.

### Face image collection

We acquired facial images from all the study participants using Samsung Galaxy A9 mobile phone (Samsung, Denmark). The mobile is fitted with a 24mp camera with capability to self-adjust brightness, color, and contrast to produce high dynamic range (HDR) images. Up to ten colored, 3X4 sized HDR images were captured at white background from each participant. After acquisition, the images were checked for their quality, and bad quality images such as with unintended brightness, contrast or coloration or images that do not meet the requirements for image analysis by artificial intelligence tools were discarded. Correctly acquired images were annotated with location and participant information to ensure where and from whom they were collected. Subsequently, the images were transferred to and stored on password protected personal computers and electronic storage devices. After we completed image collection from all the study participants, we performed de-identification by removing all the identifiers of the participants and the images were coded to ensure confidentiality.

### Face image-based age prediction

We used three different freely online available AI methods to predict age of each study participant from their facial images. These are method 1 (https://www.facialage.com/), method 2 (https://howolddoyoulook.com/) and method 3 (https://age.toolpie.com/). Each method is built-in with algorithms to perform a step-by-step analysis regarding the uploaded image quality and features in the face skin to produce estimated age. Using each predictor, we performed three independent analyses on three different face images of each participant and produced three independent predicted ages per participant. The predicted ages of each participant from the three methods were averaged and the mean predicted age of each participant was used in further analysis.

### Thin blood smear collection and staining

After we captured face images, we subsequently collected peripheral blood smears from each highland and lowland participant as well. Up to five thin blood smears per participant were prepared from the middle fingertip by finger prick. Air-dried smears were fixed in absolute methanol and stained in freshly prepared Giemsa stain as previously described ^42^. To avoid in-group and between group staining variation, both highland and lowland smears were stained in the same working Giemsa solution, simultaneously.

### Peripheral blood mononuclear cell (PBMC) image acquisition

We examined the Giemsa-stained blood films under oil immersion OPTIKA light microscope (Optika, Italy) for the detection of peripheral blood mononuclear cells (PBMCs). The microscope is fitted with a camera and linked to Optika Vision Pro version 2.7 software on a desktop where the images of PBMCs are displayed and captured. Peripheral blood mononuclear cells include monocytes and lymphocytes, and detection and identification of these cells from each other or from other blood cells was done based on cell size, nuclear shape, granulation and/ or staining properties ^42^. We captured approximately 100 images of PBMCs per participant (70% lymphocytes), and more than 43,000 images from all the study participants.

### Peripheral blood mononuclear cell (PBMC) senescence prediction

We used five different models to detect senescence in monocytes and lymphocytes of each participant. Each of these models is specialized to recognize a specific type of senescence. Notably, the RS model is specific to detect senescence due to replicative exhaustion, whereas the IR and Doxo models are trained on senescent cells from DNA damage attributed to ionizing radiation (IR) and doxorubicin (Doxo) treatment, respectively. The Anti and Atvr models detect senescence caused by antimycin-A-induced mitochondrial dysfunction and atazanavir/ ritonavir-induced proteotoxicity, respectively ^43^. More than 43,000 captured images of the PBMCs (∼70% lymphocytes) were uploaded to each model, and the model-specific algorithms analyze the nuclear morphology of the PBMCs and generated cell type-specific senescence scores for each participant ^23^.

### Statistical analysis

IBM SPSS Statistics Version 29.0.1.0 (171) for Mac OS was used to determine the correlation of risk factors and disease burden with elevation. Graphs and heatmaps showing the trend and rank of risk factors and disease burden were done in Microsoft Excel 365. All scatter plots and linked data processing, statistical analysis and graphs were made utilizing GraphPad Prism Version 10.1.1 for Mac OS (GraphPad Software, Boston, Massachusetts USA). The descriptive statistics of the continuous variables of the study participants were presented as mean ± standard deviation, all other variables, as counts. Delta age or age difference or aging acceleration or deceleration was calculated by subtracting the chronological age from the predicted age. Simple linear regression and Pearson’s correlation was applied to determine all the relationship studies. In all analyses, significance was considered at p-values less than 0.05.

### Ethical permission

This study was based on data from human participants and public databases. All human studies were performed after gaining ethical approval from the Institutional Review Board of the College of Health Sciences at Mekelle University (MU-IRB 2020/2022) in Ethiopia. We collected samples and covariate data after gathering informed consent from all participants. We adopted guidelines from local sources and the Danish Code of Conduct for Research Integrity and conducted the study in accordance with the Declaration of Helsinki. Since our study involved personal data, we performed data anonymization and then coding before any data analysis or sharing was made.

## Data availability statement

Data not in this article are included as supplementary material. If further data is required, inquiries can be forwarded to the corresponding author.

## Author Contributions

AT conceived, designed, collected and processed the samples, analyzed the data, and wrote the manuscript. IH developed deep learning models, generated senescence data and edited the manuscript; MP edited the manuscript; DB edited the manuscript; GK edited the manuscript; MSK conceived, designed, supervised the project and edited the manuscript.

## Roel of the funding source

We declare that the funders had no role in the scientific aspect of this study including in the study design, data collection and analysis, decision to publish, or manuscript preparation.

## Data Availability

Non-person identifiable data is available upon reasonable request.

## Acknowledgments

This research was supported by the Novo Nordisk Foundation Challenge Programme (NNF17OC0027812), the Nordea Foundation (02-2017-1749), the Neye Foundation, the Lundbeck Foundation (R324-2019-1492), the Ministry of Higher Education and Science (0238- 00003B), VitaDAO, and Insilico Medicine.

We would like to acknowledge the Tigray Statistics Agency, specially, Halefom Hagos for making the Ethiopian and Tigray maps as well as generating the elevation of the tabiyas where the study participants reside.

This study involved participants from lowland and highlands of the Tigray region in Northern Ethiopia, and we thank for their volunteer participation and providing their facial images, blood samples and covariate data. We also thank the Tigray Health Bureau and the health facilities under it for providing facility during sample and data collection. We extend our appreciation to the Pathology Unit of the College of Health Sciences at Mekelle University for providing staining and imaging facilities.

---

**Figure S1:** **Study area and setting to investigate exposome, disease burden and human aging at low and high altitude regions in Ethiopia.** a. Elevation map of Ethiopia and its subnational regions. b. Mean elevation and elevation-based changes in the partial pressure of oxygen in the inspired air of the subnational regions of Ethiopia. c. Facial images and blood smears collection sites in Tigray. d. Mean elevation of the sample collection sites with estimated inhaled oxygen concentration.

**Figure S2:** Demographic features of the Tigray population. a. Total population by gender at five years interval from 2007 to 2021. b. Percent increase of population from 2007 to 2021. c. Total population in rural and urban at five years interval from 2007 to 2021. d. Age aggregated population size by year. e. Age aggregated population size by gender. f. Age aggregated population size by residency. g. Percent of population by sex at 500 masl elevation interval. h. Percent of population by residency at 500 masl elevation interval.

**Figure S3:** Variation and relatedness of age predictor methods from facial images. a. PCA or predicted age by the four different age predictors b. Correlation result of the predicted age by the four different age predictors

**Figure S4:** Predicted age from facial images of highland and lowland dwellers in Tigray. a. Scatter plot of averaged predicted age with the residential elevation of the participants. b. Scatter plot of averaged predicted age with the chronological age of the participants. c. Scatter plot of averaged predicted age with the body mass index of the participants. d. Scatter plot of averaged predicted age with the gender of the participants. e. Scatter plot of averaged predicted age with the residence of the participants. f. Scatter plot of predicted age by method 1 with the residential elevation of the participants. g. Scatter plot of predicted age by method 1 with the chronological age of the participants. h. Scatter plot of predicted age by method 1 with the body mass index of the participants. i. Scatter plot of predicted age by method 1 with the gender of the participants. j. Scatter plot of predicted age by method 1 with the residence of the participants. k. Scatter plot of predicted age by method 2 with the residential elevation of the participants. l. Scatter plot of predicted age by method 2 with the chronological age of the participants. m. Scatter plot of predicted age by method 2 with the body mass index of the participants. n. Scatter plot of predicted age by method 2 with the gender of the participants. o. Scatter plot of predicted age by method 2 with the residence of the participants. p. Scatter plot of predicted age by method 3 with the residential elevation of the participants. q. Scatter plot of predicted age by method 3 with the chronological age of the participants. r. Scatter plot of predicted age by method 3 with the body mass index of the participants. s. Scatter plot of predicted age by method 3 with the gender of the participants. t. Scatter plot of predicted age by method 3 with the residence of the participants.

**Figure S5:** Senescence status of highland and lowland dwellers. a. Scatter plot of predicted senescence (RS model) of monocytes and lymphocytes with the body mass index of the study participants. b. Scatter plot of predicted senescence (RS model) of monocytes and lymphocytes with the gender of the study participants. c. Scatter plot of predicted senescence (RS model) of monocytes and lymphocytes with the residence of the study participants. d. Scatter plot of predicted senescence (IR model) of monocytes and lymphocytes with the body mass index of the study participants. e. Scatter plot of predicted senescence (IR model) of monocytes and lymphocytes with the gender of the study participants. f. Scatter plot of predicted senescence (IR model) of monocytes and lymphocytes with the residence of the study participants. g. Scatter plot of predicted senescence (Doxo model) of monocytes and lymphocytes with the body mass index of the study participants. h. Scatter plot of predicted senescence (Doxo model) of monocytes and lymphocytes with the gender of the study participants. i. Scatter plot of predicted senescence (Doxo model) of monocytes and lymphocytes with the residence of the study participants. j. Scatter plot of predicted senescence (Atvr model) of monocytes and lymphocytes with the body mass index of the study participants. k. Scatter plot of predicted senescence (Atvr model) of monocytes and lymphocytes with the gender of the study participants. l. Scatter plot of predicted senescence (Atvr model) of monocytes and lymphocytes with the residence of the study participants. m. Scatter plot of predicted senescence (Anti model) of monocytes and lymphocytes with the body mass index of the study participants. n. Scatter plot of predicted senescence (Anti model) of monocytes and lymphocytes with the gender of the study participants. o. Scatter plot of predicted senescence (Anti model) of monocytes and lymphocytes with the residence of the study participants.

