## Supplemental data for "Deep learning reveals diverging effects of altitude on aging"

[3]Tracked Biotechnologies

**Contains:**

Supplemental figures 1-5

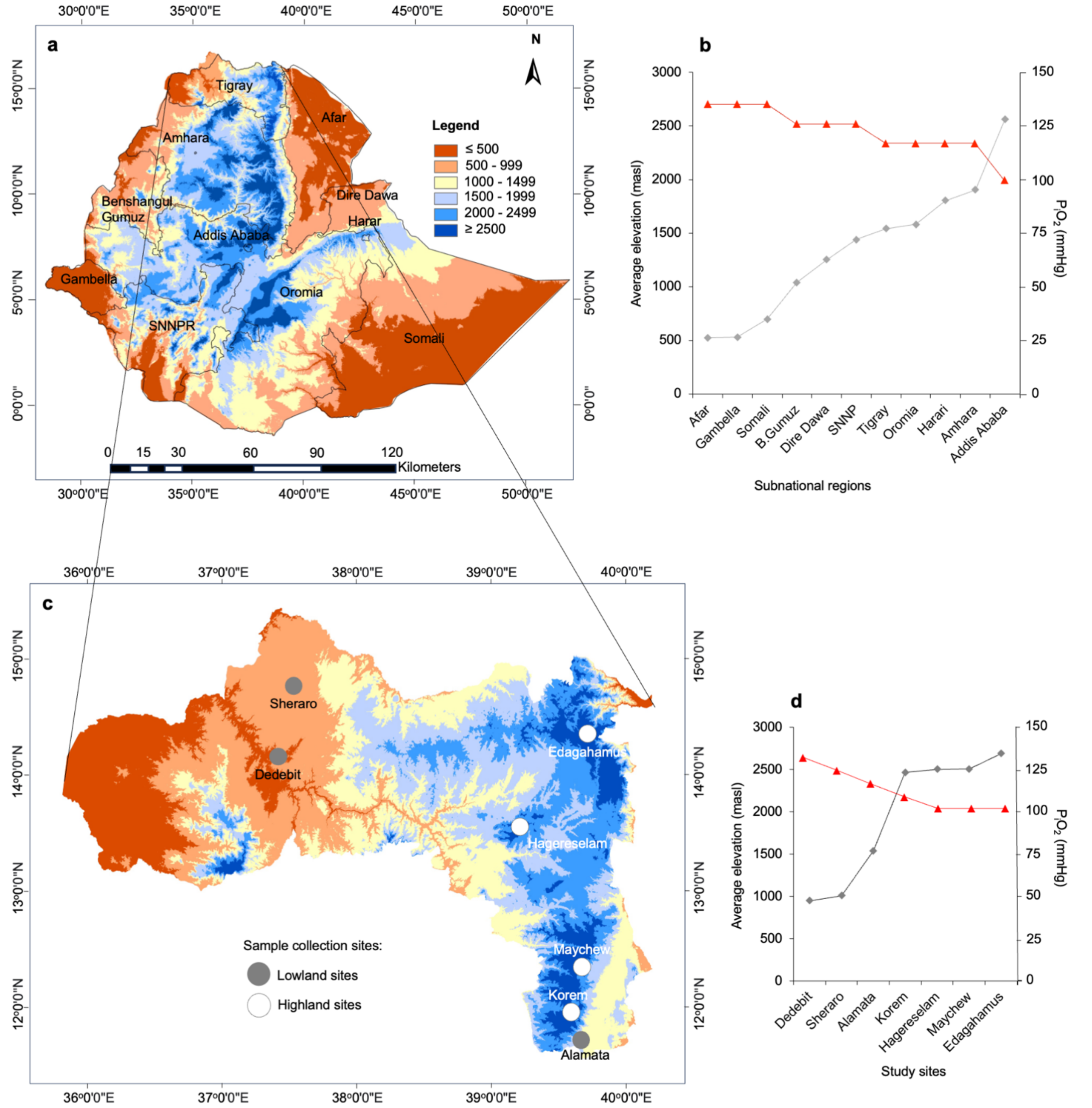


**Figure S1: Study area and setting to investigate human aging at high altitude. a** Elevation map of Ethiopia and its subnational regions. **b** Mean elevation and elevation-based changes in the partial pressure of oxygen in the inspired air of the subnational regions of Ethiopia. **c** Facial images and blood smears collection sites in Tigray. **d** Mean elevation of the sample collection sites with estimated inhaled oxygen concentration.

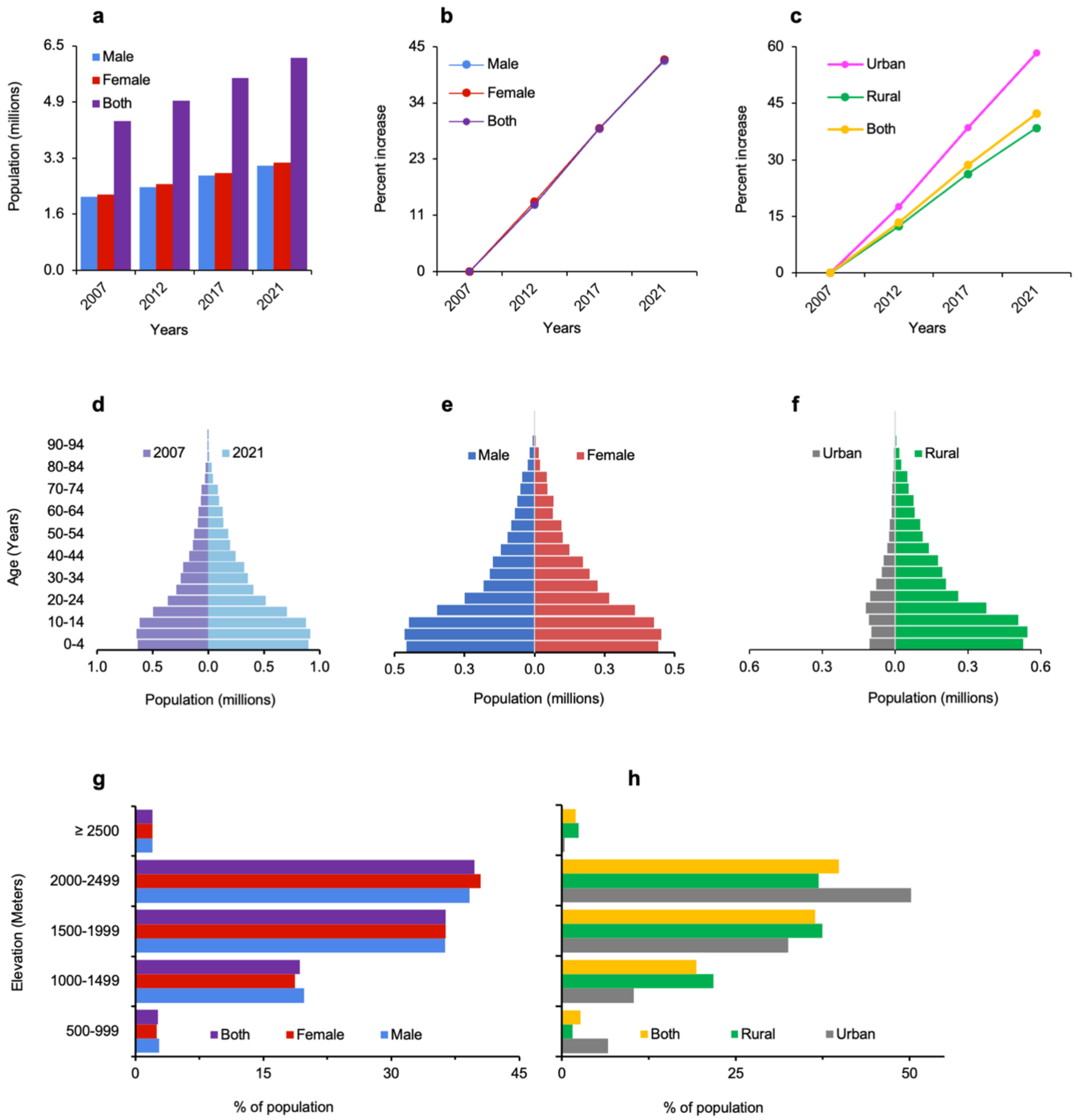


**Figure S2: Demographic features of the Tigray population. a** Total population by gender at five years interval from 2007 to 2021. **b** Percent increase of population from 2007 to 2021. **c** Total population in rural and urban at five years interval from 2007 to 2021. **d** Age aggregated population size by year. **e** Age aggregated population size by gender. **f** Age aggregated population size by residency. **g** Percent of population by sex at 500 masl elevation interval. **h** Percent of population by residency at 500 masl elevation interval.

**Age predictor methods from facial images**

Four methods applied:

Method 1 (PC1): https://saas.haut.ai/
Method 2 (PC2): https://www.facialage.com/
Method 3 (PC3): https://howolddoyoulook.com/
Method 4 (PC4): https://age.toolpie.com/

**Variation and relatedness of the methods in age prediction**

**a**. PCA analysis

| | PC1 | PC2 | PC3 | PC4 |
|---|---|---|---|---|
| Eigenvalue | 3,743 | 0,1991 | 0,05619 | 0,001907 |
| Proportion of variance | 93,57% | 4,98% | 1,40% | 0,05% |
| Cumulative proportion of variance | 93,57% | 98,55% | 99,95% | 100,00% |

**b**. Correlation analysis among the predicted age by the four methods

| **Pearson r** | Hault | Face detector | Howoldyou | Toolpie |
|---|---|---|---|---|
| Hault | 1 | 0,86 | 0,85 | 0,86 |
| Face detector | 0,86 | 1 | 0,96 | 100 |
| Howoldyou | 0,85 | 0,96 | 1 | 0,96 |
| Toolpie | 0,86 | 100 | 0,96 | 1 |

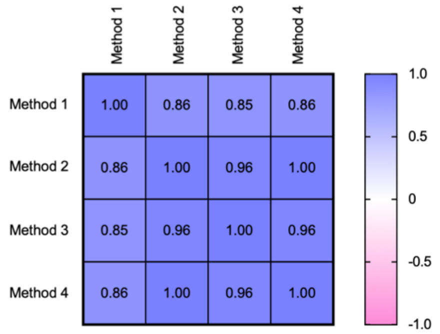


**Figure S3: Variation and relatedness of age predictor methods from facial images**. **a** PCA or predicted age by the four different age predictors. **b** Correlation result of the predicted age by the four different age predictors.

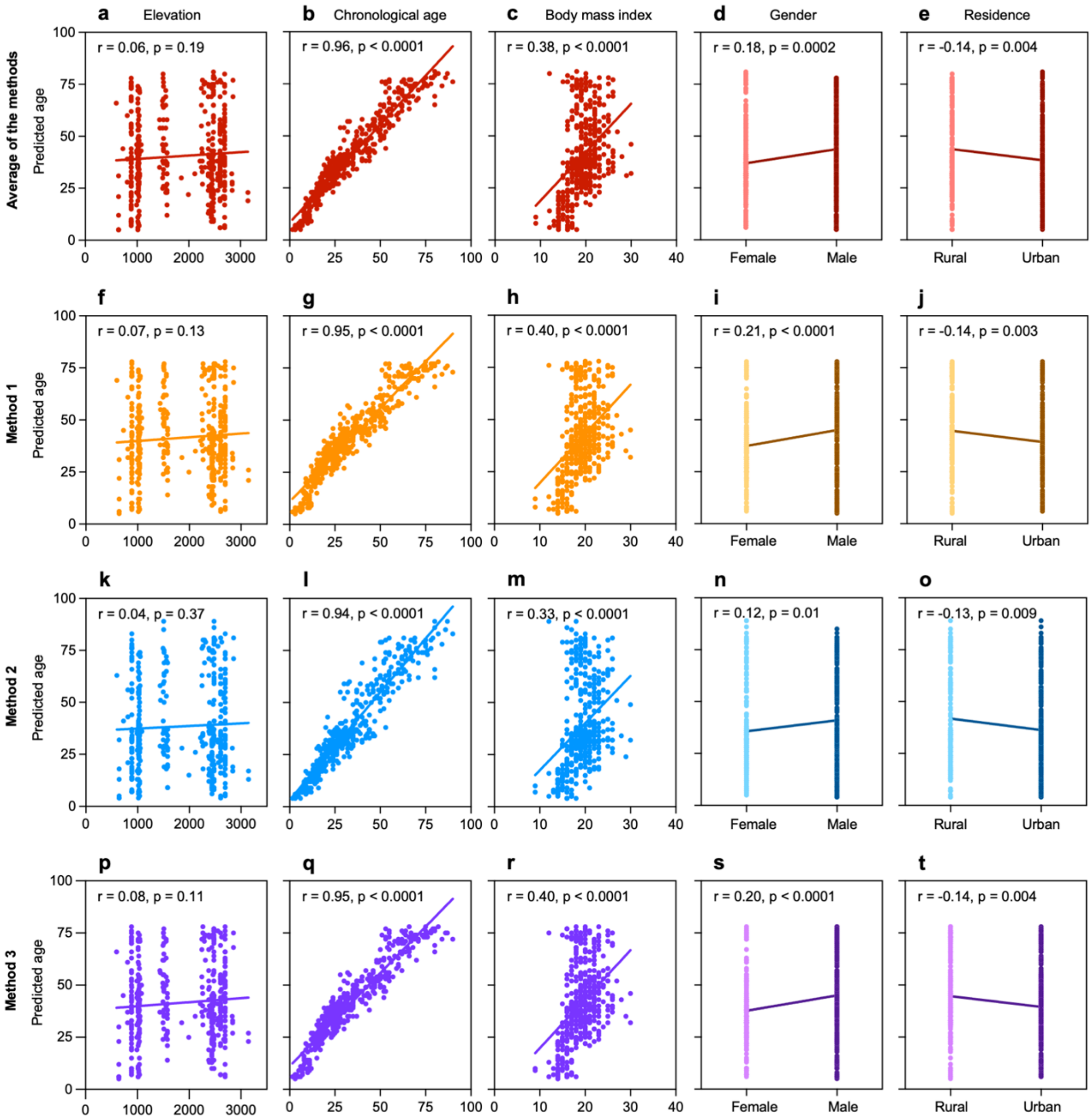


**Figure S4: Predicted age from facial images of highland and lowland dwellers in Tigray.** **a** Scatter plot of averaged predicted age with the residential elevation of the participants. **b** Scatter plot of averaged predicted age with the chronological age of the participants. **c** Scatter plot of averaged predicted age with the body mass index of the participants. **d** Scatter plot of averaged predicted age with the gender of the participants. **e** Scatter plot of averaged predicted age with the residence of the participants. **f** Scatter plot of predicted age by method 1 with the residential elevation of the participants. **g** Scatter plot of predicted age by method 1 with the chronological age of the participants. **h** Scatter plot of predicted age by method 1 with the body mass index of the participants. **i** Scatter plot of predicted age by method 1 with the gender of the participants. **j** Scatter plot of predicted age by method 1 with the residence of the participants. **k** Scatter plot of predicted age by method 2 with the residential elevation of the participants. **l** Scatter plot of predicted age by method 2 with the chronological age of the participants. **m** Scatter plot of predicted age by method 2 with the body mass index of the participants. **n** Scatter plot of predicted age by method 2 with the gender of the participants. **o** Scatter plot of predicted age by method 2 with the residence of the participants. **p** Scatter plot of predicted age by method 3 with the residential elevation of the participants. **q** Scatter plot of predicted age by method 3 with the chronological age of the participants. **r** Scatter plot of predicted age by method 3 with the body mass index of the participants. **s** Scatter plot of predicted age by method 3 with the gender of the participants. **t** Scatter plot of predicted age by method 3 with the residence of the participants.

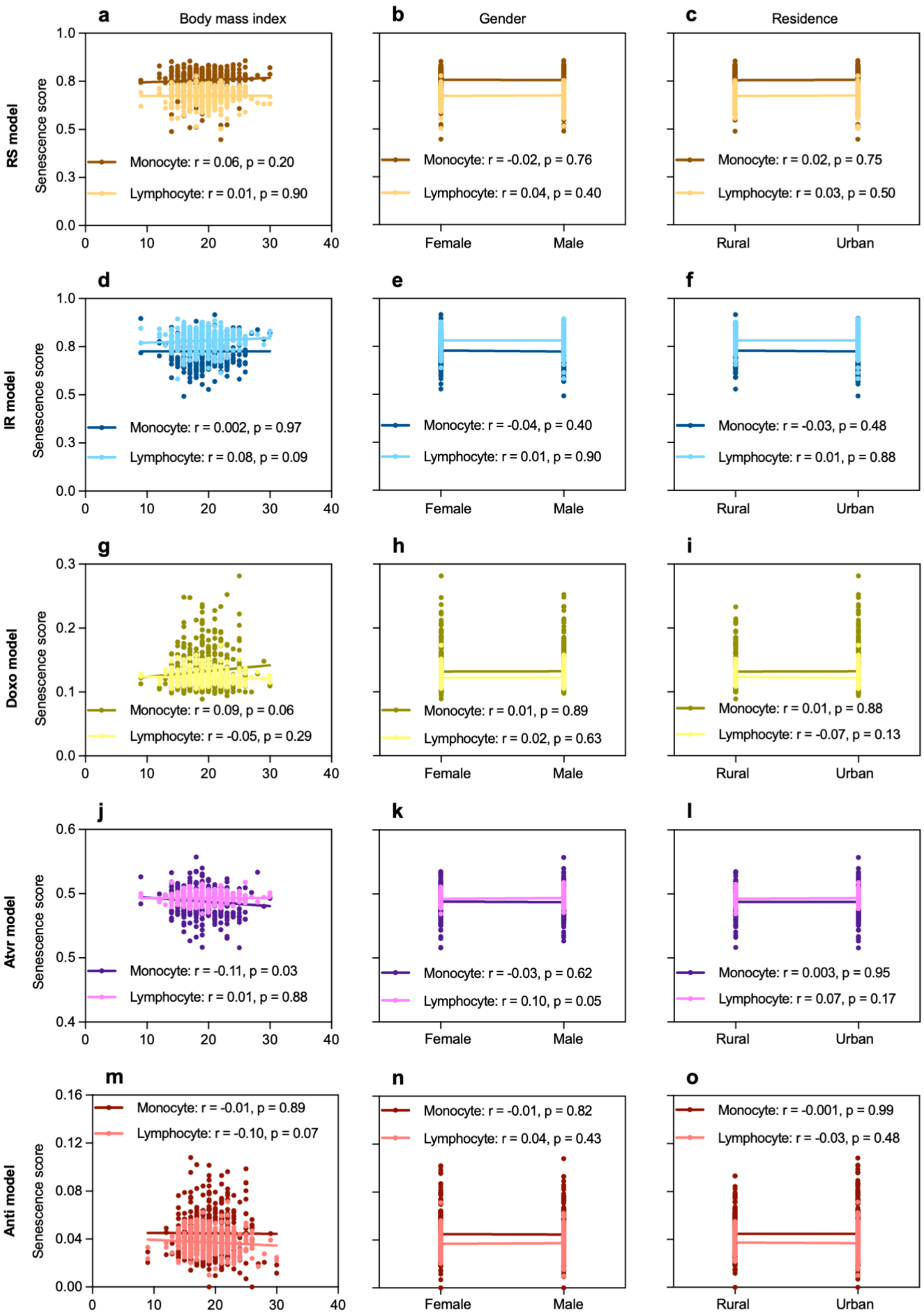

a
Body mass index
b
Gender
c
Residence
RS model
Senescence score
Monocyte: r = 0.06, p = 0.20
Lymphocyte: r = 0.01, p = 0.90
Monocyte: r = -0.02, p = 0.76
Lymphocyte: r = 0.04, p = 0.40
Monocyte: r = 0.02, p = 0.75
Lymphocyte: r = 0.03, p = 0.50
d
e
f
IR model
Monocyte: r = 0.002, p = 0.97
Lymphocyte: r = 0.08, p = 0.09
Monocyte: r = -0.04, p = 0.40
Lymphocyte: r = 0.01, p = 0.90
Monocyte: r = -0.03, p = 0.48
Lymphocyte: r = 0.01, p = 0.88
g
h
i
Doxo model
Monocyte: r = 0.09, p = 0.06
Lymphocyte: r = -0.05, p = 0.29
Monocyte: r = 0.01, p = 0.89
Lymphocyte: r = 0.02, p = 0.63
Monocyte: r = 0.01, p = 0.88
Lymphocyte: r = -0.07, p = 0.13
j
k
l
Atvr model
Monocyte: r = -0.11, p = 0.03
Lymphocyte: r = 0.01, p = 0.88
Monocyte: r = -0.03, p = 0.62
Lymphocyte: r = 0.10, p = 0.05
Monocyte: r = 0.003, p = 0.95
Lymphocyte: r = 0.07, p = 0.17
m
n
o
Anti model
Monocyte: r = -0.01, p = 0.89
Lymphocyte: r = -0.10, p = 0.07
Monocyte: r = -0.01, p = 0.82
Lymphocyte: r = 0.04, p = 0.43
Monocyte: r = -0.001, p = 0.99
Lymphocyte: r = -0.03, p = 0.48
Female
Male
Rural
Urban
